# Viral load and infectivity of SARS-CoV-2 in paired respiratory and oral specimens from symptomatic, asymptomatic or post-symptomatic individuals

**DOI:** 10.1101/2021.11.13.21266305

**Authors:** Rebecca L. Tallmadge, Melissa Laverack, Brittany Cronk, Roopa Venugopalan, Mathias Martins, XiuLin Zhang, François Elvinger, Elizabeth Plocharczyk, Diego G. Diel

## Abstract

In the present study, we assessed the diagnostic sensitivity and determined the viral load and infectivity of SARS-CoV-2 in paired respiratory (nasopharyngeal and anterior nares) and oral samples (saliva and sublingual swab). Samples were collected from 77 individuals of which 75 were diagnosed with COVID-19 and classified as symptomatic (n=29), asymptomatic (n=31), or post-symptomatic (n=15). Specimens were collected at one time point from each individual, between day 1 to 23 after the initial COVID-19 diagnosis, and included self-collected saliva (S), or sublingual (SL) swab, and bilateral anterior nares (AN) swab, followed by healthcare provider collected nasopharyngeal (NP) swab. Sixty-three specimen sets were tested using five assay/platforms. The diagnostic sensitivity of each assay/platform and specimen type was determined. Of the 63 specimen sets, SARS-CoV-2 was detected in 62 NP specimens, 52 AN specimens, 59 saliva specimens, and 31 SL specimens by at least one platform. Infectious SARS-CoV-2 was isolated from 21 NP, 13 AN, 12 saliva, and one SL specimen out of 50 specimen sets. SARS-CoV-2 isolation was most successful up to 5 days after initial COVID-19 diagnosis using NP specimens from symptomatic patients (16 of 24 positives, 66.67%), followed by specimens from asymptomatic patients (5 of 17 positives, 29.41%), while it was not very successful with specimens from post-symptomatic patients. Benefits of self-collected saliva and AN specimens balance the loss of sensitivity relative to NP specimens. Therefore, saliva and AN specimens are acceptable alternatives for symptomatic SARS-CoV-2 diagnostic testing or surveillance with increased sampling frequency of asymptomatic individuals.

**Importance:** The dynamics of infection with SARS-CoV-2 has a significant impact on virus infectivity and in the diagnostic sensitivity of molecular and classic virus detection tests. In the present study we determined the diagnostic sensitivity of paired respiratory (nasopharyngeal and anterior nares swabs) and oral secretions (saliva and sublingual swab) and assessed infectious virus shedding patterns by symptomatic, asymptomatic or post-symptomatic individuals. Understanding the diagnostic performance of these specimens and the patterns of infectious virus shedding in these bodily secretions provides critical information to control COVID-19, and may help to refine guidelines on isolation and quarantine of positive individuals and their close contacts identified through epidemiological investigations.

## Introduction

The emergence of the severe acute respiratory syndrome coronavirus 2 (SARS-CoV-2), causative agent of coronavirus disease 19 (COVID-19), in December 2019 led to an unprecedented pandemic that has killed and continues to kill millions of people worldwide (1). The number of COVID-19 cases increased rapidly since the identification of the virus in Wuhan, China. The virus presents a basic reproductive rate estimated at 2.2 – 2.68 and an epidemic doubling time of 6.4 days (2, 3). Retrospective studies indicate that individuals infected with SARS-CoV-2 and exhibiting symptoms are infectious for approximately 9 days, although the presence of viral RNA can linger beyond the end of the infectious period (4). An important characteristic that favors the efficient spread of SARS-CoV-2 is the fact that virus shedding occurs prior to the onset of symptoms, and it has been estimated that ∼44% of the infections occur while the index case is pre-symptomatic (5, 6). To date, over 247 million COVID-19 cases have been confirmed across the globe and more than 5 million of these cases have resulted in death (https://covid19.who.int/, accessed on 11/4/2021). Although vaccines are now available, the virus continues to cause a toll to human health and several countries are undergoing additional epidemic waves leading to significant public health concerns; especially due to reluctance of a great proportion of the population to vaccination, evidence of incomplete protection from vaccination, and the emergence of SARS-CoV-2 variants (7–12). Thus, the demand for rapid, sensitive, and efficient diagnostic tests remains. A refined understanding of the SARS-CoV-2 infectious period is also needed for intervention to limit transmission.

Transmission of SARS-CoV-2 occurs mainly via aerosols and droplets and infection can cause broad clinical symptoms in affected individuals (13, https://www.cdc.gov/coronavirus/2019-ncov/symptoms-testing/symptoms.html). Because of the respiratory-based mode of transmission and diversity of symptoms, a wide range of specimen types have been evaluated for the detection of SARS-CoV-2 RNA (14, 15). Overall, moderate to high rates of detection were found in lower respiratory tract secretions (sputum, bronchoalveolar lavage fluid), respiratory swabs (nasopharyngeal swabs, nasal swabs, throat swabs, pharyngeal swabs, oropharyngeal swabs), saliva, feces/rectal swabs, and serum (14, 15). Blood and urine specimens provided the lowest rates of SARS-CoV-2 RNA detection (14, 15). The Centers for Disease Control and Prevention recommend an upper respiratory specimen for initial testing of a suspect SARS-CoV-2 infection, which may include a nasopharyngeal, oropharyngeal, nasal mid-turbinate, anterior nares, or saliva specimen (https://www.cdc.gov/coronavirus/2019-ncov/lab/guidelines-clinical-specimens.html). Collection of NP swabs, OP swabs, NP washes/aspirates or nasal washes/aspirates require a trained healthcare provider, whereas other upper respiratory swabs (e.g anterior nares) and saliva specimens may be self-collected, offering advantages of limiting healthcare provider exposure to the virus and reducing the need and use for personal protective equipment during collection.

In the present study, we compared the clinical performance of three diagnostic assays (Rheonix COVID-19 MDx assay, EZ-SARS-CoV-2 real-time RT-PCR assay, and the TaqPath COVID-19 Combo kit assay) and determined the diagnostic sensitivity of paired respiratory (nasopharyngeal and anterior nares swabs) and oral samples (saliva and sublingual swabs) collected from symptomatic, asymptomatic, or post-symptomatic individuals. Additionally, we determined the viral load and compared shedding of infectious virus in the different specimen types.

## Materials and Methods

### Study design and specimen collection

Seventy-seven specimen sets were collected for validation of clinical diagnostic tests under Cayuga Medical Center (CMC) Institutional Review Board approval 0420EP. The Cornell University Institutional Review Board (IRB) for Human Participants also reviewed and approved the study (protocol number 2007009706). All patients agreed to participate and provided verbal consent prior to specimen collection.

Nasopharyngeal (NP) specimens were collected by a healthcare provider with 3D-printed swabs (NP Swab O1, Origin, San Francisco, CA) (16). Anterior nares (AN) swabs were self-collected under observation. Each patient was provided with a nasal swab and instructed to insert the swab less than one inch into the anterior nostril and rotate the swab for 10 seconds against the nasal wall, then repeat the procedure in the counter lateral nostril using the same swab. Sublingual (SL) swabs were collected by instructing the patient to place the swab under the tongue and rotate it for one minute. Anterior nares and sublingual swabs were collected using either a nylon tipped sampling swab (ASP Medical Disposable sampling swab 8205, Cardinal Health, Dublin, OH), a CultureSwab™ Liquid Stuart Single Swab (220099, BD Life Sciences, Sparks, MD), or a CultureSwab™ Liquid Stuart Double Swab (220109, BD Life Sciences, Sparks, MD). All swabs were immersed in 800 μl viral transport media immediately after collection and stored under refrigeration. For saliva (S) collection, patients were instructed not to eat, drink, or chew gum or tobacco for at least 30 minutes before sampling. Patients were instructed to drool 3 mL of saliva into a 15 mL conical tube. An inactivating medium containing guanidine hydrochloride was added to the saliva and mixed by shaking the tightly closed tube. Each specimen was collected in duplicate and submitted for testing at the CMC testing laboratory and at the Cornell COVID-19 Testing Laboratory (CCTL).

Patients were classified as symptomatic (n=29) if they had a positive COVID-19 diagnostic test within 5 days of sampling and they exhibited any of the following symptoms: fever, chills, dyspnea, nausea, vomiting, diarrhea, cough, sore throat, fatigue, muscle aches, congestion, runny nose, new loss of taste or smell. Patients were classified as asymptomatic (n=31) if they had a positive COVID-19 diagnostic test within 5 days of sampling and they did not exhibit any of the symptoms listed above. Patients were classified as post-symptomatic (n=15) if they were sampled more than 5 days after their initial positive COVID-19 RT-PCR diagnostic test and were no longer presenting symptoms at the time of re-sampling for this study. Because the asymptomatic cohort was included in this study, specimen collection days were tracked relative to the first diagnostic test confirming SARS-CoV-2 infection rather than days since symptom onset.

### Nucleic acid extraction

The MagMax Viral/Pathogen II (MVP II) Nucleic Acid Isolation Kit (Applied Biosystems, Foster City, CA) was used to extract nucleic acid (NA) from 200 μl of each specimen. A negative extraction control containing 200 μl viral transport media (Corning, Corning, NY) was included on every plate. Extractions were processed on the Kingfisher Flex Magnetic Particle Processor (Thermo Fisher Scientific Inc., Waltham, MA). The resulting elution volume was 50 μl and the same elution was used for testing with the EZ-SARS-CoV-2 RT-PCR and the TaqPath COVID-19 Combo Kit Multiplex Real-Time RT-PCR assays.

### SARS-CoV-2 Detection assays

The Rheonix COVID-19™ MDx assay (Rheonix Inc., Ithaca, NY) is an automated endpoint RT-PCR assay that has an Emergency Use Authorization (EUA) from the U. S. Food and Drug Administration (FDA). The fully automated Rheonix Encompass MDx^®^ workstation was used as recommended by the manufacturer for this work at the Cayuga Medical Center, Ithaca, NY. The Rheonix assay targets the N gene of the SARS-CoV-2 genome and includes detection of the human RNase P gene as an internal control. Approximately 0.5 ml of each specimen type was loaded in the Rheonix Encompass MDx^®^ reaction tube and tested. Negative and positive controls were included in each run. An “error” result indicated that an error occurred during the run which prevented a valid result interpretation.

The EZ-SARS-CoV-2 Real-Time RT-PCR assay (Tetracore, Inc., Rockville, MD) was performed as indicated by the manufacturer. The EZ-SARS-CoV-2 RT-PCR assay was validated for NP and AN specimens (17). A positive amplification control provided by the manufacturer and a negative amplification control was included on every plate.

The Thermo Fisher TaqPath COVID-19 Combo Kit Multiplex Real-Time RT-PCR assay (Thermo Fisher Scientific Inc.) was performed and analyzed as directed by the manufacturer. COVID-19 Interpretive Software version 1.2 (Applied Biosystems, Foster City, CA) was used to interpret the results. An “inconclusive” result indicated that only one of the three SARS-CoV-2 targets were detected. This assay is intended for use with NP and AN swabs under an EUA from the FDA. A positive amplification control provided by the manufacturer and a negative amplification control was included on every plate.

Real-time PCR assays were performed on both ABI 7500 Fast and QuantStudio 5 real-time PCR instruments (Thermo Fisher Scientific Inc.), using cycling parameters recommended by the respective manufacturer.

### Diagnostic sensitivity

The diagnostic sensitivity was calculated for each platform by dividing the number of positive samples within a specimen type by the total number of infected patients.

### Virus isolation

Virus isolation was performed in NP, AN, saliva and sublingual swab samples under biosafety level 3 (BSL-3) conditions. For this, twenty-four well plates were seeded with ∼75,000 Vero E6/TMPRSS2 cells per well 24 h prior to sample inoculation. Cells were rinsed with phosphate buffered saline (PBS) (Corning^®^) and inoculated with 150 μl of each sample and inoculum adsorbed for 1 h at 37□°C with 5% CO_2_. Mock-inoculated cells were used as negative controls. After adsorption, replacement cell culture media (Dulbecco’s modified eagle medium (DMEM) supplemented with 10% fetal bovine serum (FBS), l-glutamine (2□mM), penicillin (100□U·ml^−1^), streptomycin (100□μg·ml^−1^), and gentamicin (50□μg·ml^−1^)) was added and cells were incubated at 37 °C with 5% CO_2_ and monitored daily for cytopathic effect (CPE) for 3 days. Cell cultures with no CPE were frozen, thawed, and subjected to two additional blind passages/inoculations in Vero E6/TMPRSS2 cell cultures. CPE positive wells and wells with no CPE at the end of the third passage, were subjected to immunofluorescence staining using SARS-CoV-2 N-specific mAbs as previously described (18).

### Statistical analysis

Diagnostic sensitivity and result concordance across specimens and platforms were calculated with Microsoft Excel (Microsoft Office Professional Plus 2019). GraphPad Prism (version 9.0.2 for Windows, GraphPad Software, San Diego, CA, USA, www.graphpad.com) was used to perform testing for normal distribution of cycle threshold (Ct) values with Shapiro-Wilk tests, comparison of Ct values using Wilcoxon matched-pairs signed rank tests or Mann-Whitney tests for unpaired comparisons, and to generate plots.

## Results

### Sample workflow and categorization

The clinical performance of three molecular SARS-CoV-2 assays was evaluated on multiple specimen sets (NP, AN, S, and SL) collected from 77 patients. Specimens were obtained in duplicate from 75 patients that had previously tested positive for SARS-CoV-2 and from two patients with previous negative SARS-CoV-2 test results (**Fig. 1**). Of the 75 specimen sets collected from patients diagnosed with COVID-19, 68 sets were tested on all 5 platforms under evaluation: Rheonix, the EZ-SARS-CoV-2 assay on two real-time RT-PCR detection systems (ABI 7500 and QuantStudio 5), and the TaqPath COVID-19 Combo kit on the same two real-time RT-PCR detection systems (ABI 7500 and QuantStudio 5). SARS-CoV-2 was detected by at least one assay/platform in 63 of the 68 sample sets. The five sample sets in which SARS-CoV-2 was not detected were excluded from further analyses as shown in **Fig. 1**. Virus isolation was performed on 50 of the 63 paired specimen sets. This experimental design allowed comparison of the diagnostic sensitivity among all specimen types and detection assays.

**Figure 1.**
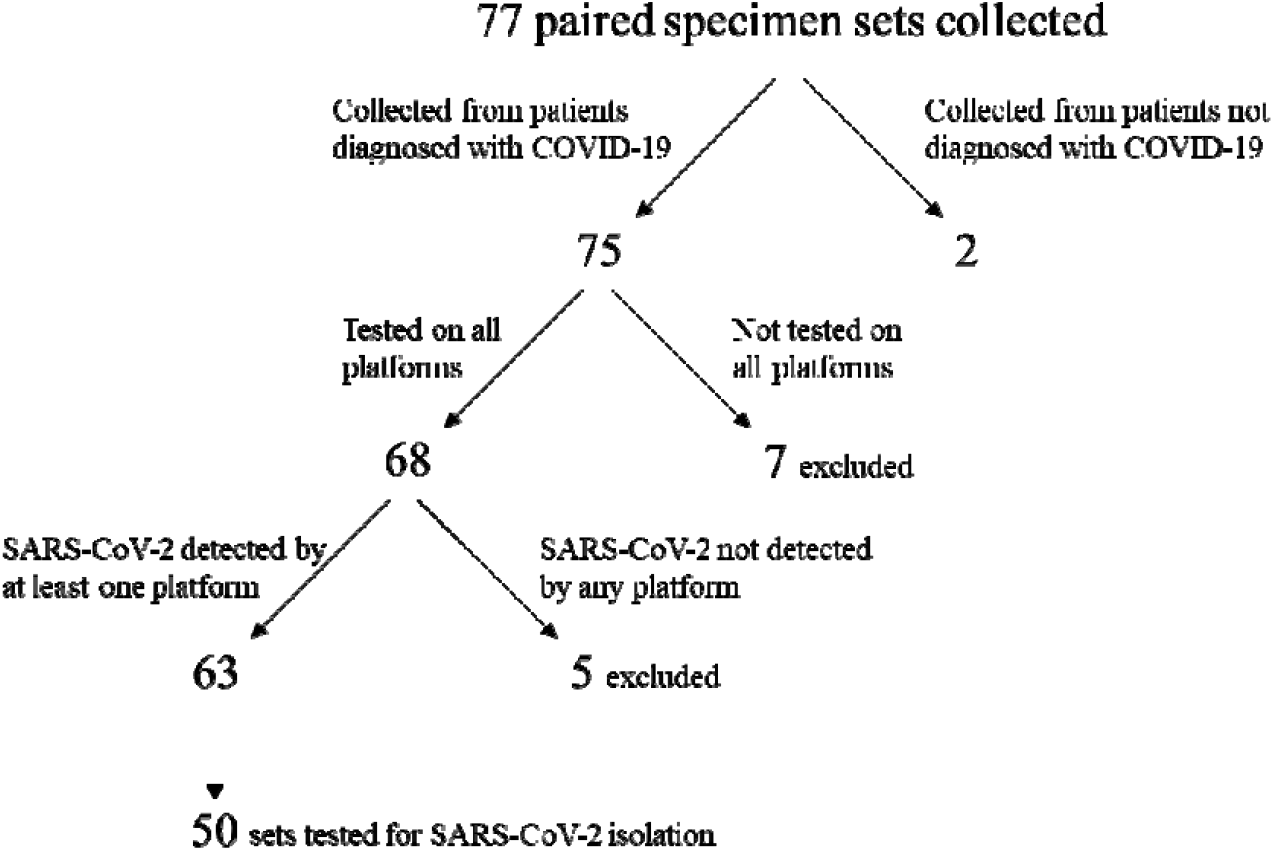
Paired specimen sets collected for this study. Specimens were excluded if testing was not performed on all platforms and if SARS-CoV-2 RNA was not detected from any specimen by any platform. After filtering, 63 sets of paired specimens were used for analyses. Of those, 50 sample sets were used for virus isolation.

To facilitate the interpretation of the results in the context of SARS-CoV-2 infection dynamics, the 63 patients included in our study were further categorized as symptomatic, asymptomatic, or post-symptomatic following the criteria described in the Materials and Methods. The SARS-CoV-2 RT-PCR results obtained from the four specimen types tested across the three assays and platforms under evaluation are shown for 25 symptomatic patients (**Table 1**), 24 asymptomatic patients (**Table 2**), and 14 post-symptomatic patients (**Table 3**). These individuals had their initial diagnostic test performed 1-23 days before the multipls specimen set was collected for this study.

**Table 1.**
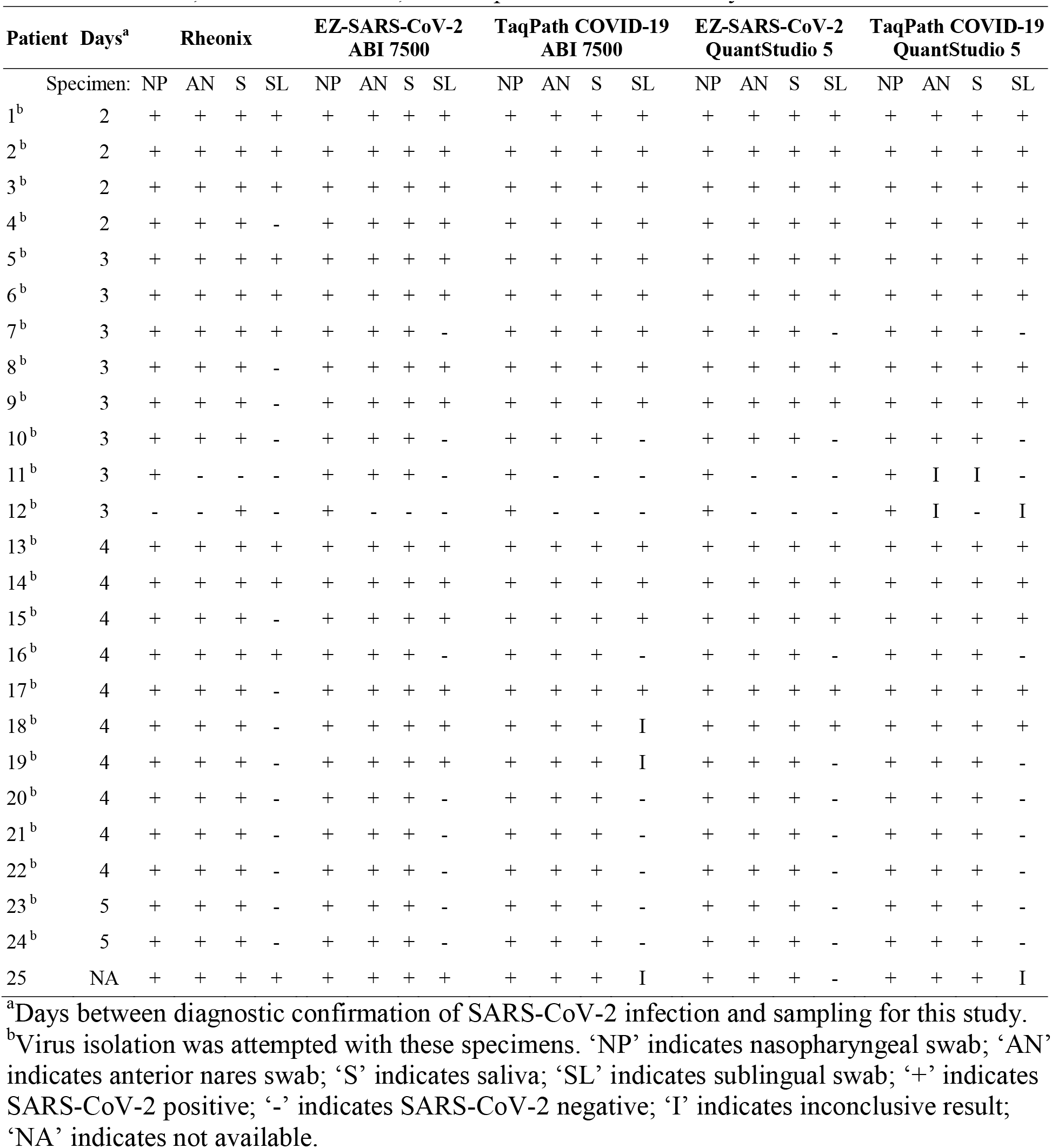
Summary of results on paired specimens collected from 25 symptomatic patients tested with the Rheonix, EZ-SARS-CoV-2, and TaqPath COVID-19 assays.

| Patient | Days<sup>a</sup> | Rheonix |  |  |  | EZ-SARS-CoV-2<br>ABI 7500 |  |  |  | TaqPath COVID-19<br>ABI 7500 |  |  |  | EZ-SARS-CoV-2<br>QuantStudio 5 |  |  |  | TaqPath COVID-19<br>QuantStudio 5 |  |  |  |
| --- | --- | --- | --- | --- | --- | --- | --- | --- | --- | --- | --- | --- | --- | --- | --- | --- | --- | --- | --- | --- | --- |
|  | Specimen: | NP | AN | S | SL | NP | AN | S | SL | NP | AN | S | SL | NP | AN | S | SL | NP | AN | S | SL |
| 1<sup>b</sup> | 2 | + | + | + | + | + | + | + | + | + | + | + | + | + | + | + | + | + | + | + | + |
| 2<sup>b</sup> | 2 | + | + | + | + | + | + | + | + | + | + | + | + | + | + | + | + | + | + | + | + |
| 3<sup>b</sup> | 2 | + | + | + | + | + | + | + | + | + | + | + | + | + | + | + | + | + | + | + | + |
| 4<sup>b</sup> | 2 | + | + | + | - | + | + | + | + | + | + | + | + | + | + | + | + | + | + | + | + |
| 5<sup>b</sup> | 3 | + | + | + | + | + | + | + | + | + | + | + | + | + | + | + | + | + | + | + | + |
| 6<sup>b</sup> | 3 | + | + | + | + | + | + | + | + | + | + | + | + | + | + | + | + | + | + | + | + |
| 7<sup>b</sup> | 3 | + | + | + | + | + | + | + | - | + | + | + | + | + | + | + | - | + | + | + | - |
| 8<sup>b</sup> | 3 | + | + | + | - | + | + | + | + | + | + | + | + | + | + | + | + | + | + | + | + |
| 9<sup>b</sup> | 3 | + | + | + | - | + | + | + | + | + | + | + | + | + | + | + | + | + | + | + | + |
| 10<sup>b</sup> | 3 | + | + | + | - | + | + | + | - | + | + | + | - | + | + | + | - | + | + | + | - |
| 11<sup>b</sup> | 3 | + | - | - | - | + | + | + | - | + | - | - | - | + | - | - | - | + | I | I | - |
| 12<sup>b</sup> | 3 | - | - | + | - | + | - | - | - | + | - | - | - | + | - | - | - | + | I | - | I |
| 13<sup>b</sup> | 4 | + | + | + | + | + | + | + | + | + | + | + | + | + | + | + | + | + | + | + | + |
| 14<sup>b</sup> | 4 | + | + | + | + | + | + | + | + | + | + | + | + | + | + | + | + | + | + | + | + |
| 15<sup>b</sup> | 4 | + | + | + | - | + | + | + | + | + | + | + | + | + | + | + | + | + | + | + | + |
| 16<sup>b</sup> | 4 | + | + | + | + | + | + | + | - | + | + | + | - | + | + | + | - | + | + | + | - |
| 17<sup>b</sup> | 4 | + | + | + | - | + | + | + | + | + | + | + | + | + | + | + | + | + | + | + | + |
| 18<sup>b</sup> | 4 | + | + | + | - | + | + | + | + | + | + | + | I | + | + | + | + | + | + | + | + |
| 19<sup>b</sup> | 4 | + | + | + | - | + | + | + | + | + | + | + | I | + | + | + | - | + | + | + | - |
| 20<sup>b</sup> | 4 | + | + | + | - | + | + | + | - | + | + | + | - | + | + | + | - | + | + | + | - |
| 21<sup>b</sup> | 4 | + | + | + | - | + | + | + | - | + | + | + | - | + | + | + | - | + | + | + | - |
| 22<sup>b</sup> | 4 | + | + | + | - | + | + | + | - | + | + | + | - | + | + | + | - | + | + | + | - |
| 23<sup>b</sup> | 5 | + | + | + | - | + | + | + | - | + | + | + | - | + | + | + | - | + | + | + | - |
| 24<sup>b</sup> | 5 | + | + | + | - | + | + | + | - | + | + | + | - | + | + | + | - | + | + | + | - |
| 25 | NA | + | + | + | + | + | + | + | + | + | + | + | I | + | + | + | - | + | + | + | I |

<sup>a</sup>Days between diagnostic confirmation of SARS-CoV-2 infection and sampling for this study.
<sup>b</sup>Virus isolation was attempted with these specimens. ‘NP’ indicates nasopharyngeal swab; ‘AN’ indicates anterior nares swab; ‘S’ indicates saliva; ‘SL’ indicates sublingual swab; ‘+’ indicates SARS-CoV-2 positive; ‘-’ indicates SARS-CoV-2 negative; ‘I’ indicates inconclusive result; ‘NA’ indicates not available.

**Table 2.** Summary of results on paired specimens collected from 24 asymptomatic patients tested with the Rheonix, EZ-SARS-CoV-2, and TaqPath COVID-19 assays.

| Patient | Days<sup>a</sup> | Rheonix |  |  |  | EZ-SARS-CoV-2<br>ABI 7500 |  |  |  | TaqPath COVID-19<br>ABI 7500 |  |  |  | EZ-SARS-CoV-2<br>QuantStudio 5 |  |  |  | TaqPath COVID-19<br>QuantStudio 5 |  |  |  |
| --- | --- | --- | --- | --- | --- | --- | --- | --- | --- | --- | --- | --- | --- | --- | --- | --- | --- | --- | --- | --- | --- |
|  | Specimen: | NP | AN | S | SL | NP | AN | S | SL | NP | AN | S | SL | NP | AN | S | SL | NP | AN | S | SL |
| 26 | 1 | + | + | + | + | + | + | + | + | + | + | + | + | + | + | + | + | + | + | + | + |
| 27 | 2 | + | + | + | + | + | + | + | + | + | + | + | + | + | + | + | + | + | + | + | + |
| 28 | 2 | + | + | + | + | + | + | + | + | + | + | + | + | + | + | + | + | + | + | + | + |
| 29<sup>b</sup> | 2 | + | + | + | - | + | + | + | - | + | + | + | I | + | + | + | - | + | + | + | I |
| 30<sup>b</sup> | 2 | + | + | + | I | + | + | + | + | + | + | - | + | + | + | - | + | + | + | I | + |
| 31<sup>b</sup> | 2 | + | + | + | I | + | + | + | - | + | + | I | + | + | + | + | - | + | + | I | + |
| 32 | 2 | + | - | I | - | - | - | - | - | - | + | - | - | - | - | - | - | - | - | - | - |
| 33 | 2 | - | - | - | - | - | - | + | + | - | - | - | - | - | - | - | - | - | - | - | - |
| 34<sup>b</sup> | 3 | + | + | + | - | + | + | + | - | + | + | + | - | + | + | + | - | + | + | + | - |
| 35<sup>b</sup> | 3 | + | + | + | - | + | + | + | - | + | + | + | - | + | + | + | - | + | + | + | - |
| 36<sup>b</sup> | 3 | + | + | + | - | + | + | + | - | + | + | + | - | + | + | + | - | + | + | + | I |
| 37<sup>b</sup> | 4 | + | + | + | + | + | + | + | + | + | + | + | + | + | + | + | + | + | + | + | + |
| 38<sup>b</sup> | 4 | + | + | + | - | + | + | + | + | + | + | + | + | + | + | + | + | + | + | + | + |
| 39<sup>b</sup> | 4 | + | + | + | - | + | + | + | - | + | + | + | - | + | + | + | - | + | + | + | - |
| 40<sup>b</sup> | 4 | + | - | + | - | + | + | + | - | + | + | + | - | + | + | + | - | + | + | + | - |
| 41<sup>b</sup> | 5 | + | + | + | + | + | + | + | + | + | + | + | I | + | + | + | + | + | + | + | + |
| 42<sup>b</sup> | 5 | + | + | + | E | + | + | + | + | + | + | + | + | + | + | + | + | + | + | + | + |
| 43<sup>b</sup> | 5 | + | + | + | - | + | + | + | - | + | + | + | - | + | + | + | - | + | + | + | - |
| 44<sup>b</sup> | 5 | + | + | + | - | + | + | + | - | + | + | + | - | + | + | + | - | + | + | + | - |
| 45<sup>b</sup> | 5 | + | + | + | - | + | + | + | - | + | + | + | - | + | + | + | - | + | + | + | - |
| 46<sup>b</sup> | 5 | + | + | + | - | + | + | + | - | + | + | + | - | + | + | + | - | + | + | + | - |
| 47<sup>b</sup> | 5 | + | + | + | - | + | + | + | - | + | + | + | - | + | + | + | - | + | + | + | - |
| 48 | NA | + | - | - | - | + | + | + | + | + | + | I | - | + | + | + | - | + | + | - | - |
| 49 | NA | + | - | - | + | + | - | - | - | + | - | - | - | + | - | - | - | + | I | - | I |

<sup>a</sup>Days between diagnostic confirmation of SARS-CoV-2 infection and sampling for this study.
<sup>b</sup>Virus isolation was attempted with these specimens. ‘NP’ indicates nasopharyngeal swab; ‘AN’ indicates anterior nares swab; ‘S’ indicates saliva; ‘SL’ indicates sublingual swab; ‘+’ indicates SARS-CoV-2 positive; ‘-’ indicates SARS-CoV-2 negative; ‘I’ indicates inconclusive result; ‘E’ indicates error; ‘NA’ indicates days not available.

**Table 3.**
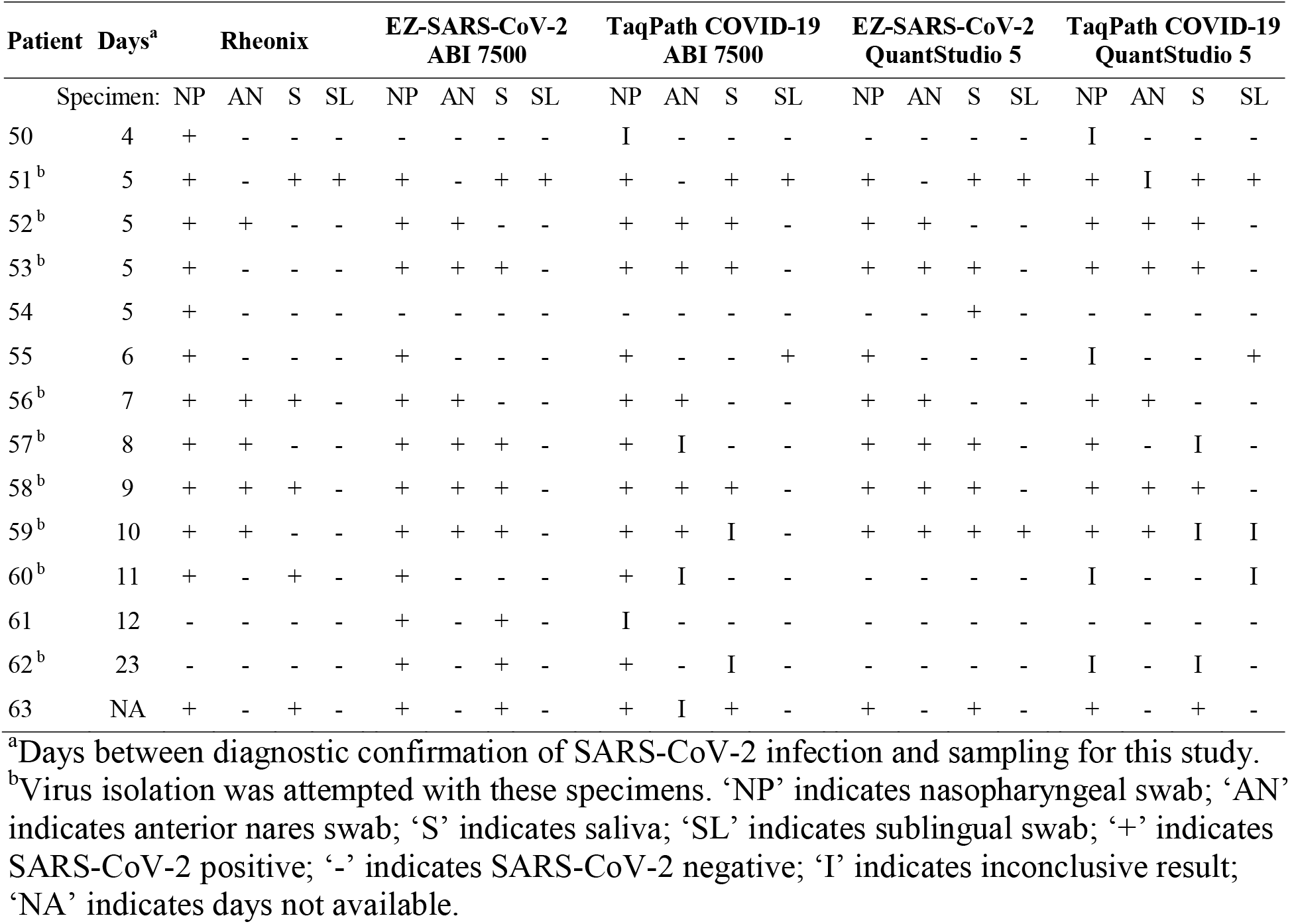
Summary of results on paired specimens collected from 14 post-symptomatic patients tested with the Rheonix, EZ-SARS-CoV-2, and TaqPath COVID-19 assays.

### Clinical performance of molecular SARS-CoV-2 assays

The diagnostic sensitivity of each specimen was evaluated across the different assays and detection platforms used in our study. The diagnostic sensitivity of each assay performed with specimens from 25 symptomatic patients is presented in **Table 4**. These specimens were collected between 2 and 5 days after the initial diagnostic test (mean of 3.42 days) and SARS-CoV-2 was detected in 96% of NP specimens on the Rheonix platform and in 100% of NP specimens with the EZ-SARS-CoV-2 or TaqPath COVID-19 real-time RT-PCR assays. SARS-CoV-2 was detected in 92 to 96% of AN and saliva specimens from symptomatic patients across all platforms. In contrast, SARS-CoV-2 detection was markedly lower in SL specimens, ranging from 40 to 60% across platforms.

**Table 4.** Comparative diagnostic sensitivity of Rheonix, EZ-SARS-CoV-2 (EZ), and TaqPath COVID-19 SARS-CoV-2 assays using paired specimens collected from 25 symptomatic patients.

| Specimen | Rheonix | EZ<br>ABI 7500 | TaqPath<br>ABI 7500 | EZ<br>QuantStudio 5 | TaqPath<br>QuantStudio 5 |
| --- | --- | --- | --- | --- | --- |
| NP | 96.00<sup>a</sup> | 100.00 | 100.00 | 100.00 | 100.00 |
| AN | 92.00 | 96.00 | 92.00 | 92.00 | 92.00 |
| S | 96.00 | 96.00 | 92.00 | 92.00 | 92.00 |
| SL | 40.00 | 60.00 | 48.00 | 56.00 | 52.00 |

<sup>a</sup>Percent diagnostic sensitivity

SARS-CoV-2 was detected by all assays and platforms in all 4 specimens from 7 of the 25 symptomatic patients. In 16 of the 25 symptomatic patients, SARS-CoV-2 was detected in NP, AN, and saliva specimens by all assays and platforms, whereas detection in SL specimens was less reliable. The remaining 2 sets of specimens were collected 3 days after the initial diagnosis, and SARS-CoV-2 was detected most consistently using NP from these patients.

The sensitivity of the assays was lower in asymptomatic patients (**Table 5**) than in symptomatic patients even though they were collected within the same time frame of 1 to 5 days after the initial diagnostic test (mean 3.41 days). Using NP specimens, the Rheonix platform detected 96% of asymptomatic patients, while EZ-SARS-CoV-2 and TaqPath COVID-19 real-time RT-PCR assays detected 92%. Detection of SARS-CoV-2 in AN specimens decreased to 79% on the Rheonix and ranged from 88 to 92% across EZ-SARS-CoV-2 and TaqPath COVID-19 real-time RT-PCR assays and platforms. The Rheonix platform presented the higher sensitivity with asymptomatic saliva specimens than AN (83%), whereas the sensitivity of the TaqPath COVID-19 assay dropped to 75%. The EZ-SARS-CoV-2 assay detected nearly 92% of saliva specimens on the ABI 7500 Fast platform, but only 83% on the QuantStudio 5 platform. Detection of SARS-CoV-2 in SL specimens was lower on all platforms, ranging from 25 to 42%.

**Table 5.** Diagnostic sensitivity of Rheonix, EZ-SARS-CoV-2 (EZ), and TaqPath COVID-19 SARS-CoV-2 assays using paired specimens collected from 24 asymptomatic patients.

| Specimen | Rheonix | EZ<br>ABI 7500 | TaqPath<br>ABI 7500 | EZ<br>QuantStudio 5 | TaqPath<br>QuantStudio 5 |
| --- | --- | --- | --- | --- | --- |
| NP | 95.83<sup>a</sup> | 91.67 | 91.67 | 91.67 | 91.67 |
| AN | 79.17 | 87.50 | 91.67 | 87.50 | 87.50 |
| S | 83.33 | 91.67 | 75.00 | 83.33 | 75.00 |
| SL | 25.00 | 41.67 | 33.33 | 33.33 | 37.50 |

<sup>a</sup>Percent diagnostic sensitivity

SARS-CoV-2 was detected in all specimen types from 4 of the 24 asymptomatic patients by all assays and platforms. Thirteen pairs of NP, AN, and saliva specimens from asymptomatic patients were detected by all assays and platforms. For some asymptomatic samples, the rate of SARS-CoV-2 detection was very low, even if they were collected 2 days after diagnosis (**Table 2**).

Diagnostic sensitivity of these SARS-CoV-2 assays using specimens from a cohort of post-symptomatic patients were lower than the symptomatic or asymptomatic patients (**Table 6**), with sample collection performed 8.46 days after initial diagnostic test on average (range 4 to 23 days). The Rheonix and EZ-SARS-CoV-2 assay on the ABI 7500 Fast platform detected nearly 86% of post-symptomatic NP specimens, although the other real-time PCR assays and platforms detected only 57 to 79% of NP specimens. Only approximately 36 to 43% of AN specimens were detected across the platforms. The EZ-SARS-CoV-2 assay was able to detect 57% of saliva specimens on the ABI 7500 Fast platform and 50% on the QuantStudio 5 platform; however, only approximately 36% were detected by the Rheonix and TaqPath COVID-19 assays. SARS-CoV-2 was only detected in 7 to 14% of post-symptomatic SL specimens.

**Table 6.** Diagnostic sensitivity of Rheonix, EZ-SARS-CoV-2 (EZ), and TaqPath COVID-19 SARS-CoV-2 assays using paired specimens collected from 14 post-symptomatic patients.

| Specimen | Rheonix | EZ<br>ABI 7500 | TaqPath<br>ABI 7500 | EZ<br>QuantStudio 5 | TaqPath<br>QuantStudio 5 |
| --- | --- | --- | --- | --- | --- |
| NP | 85.71<sup>a</sup> | 85.71 | 78.57 | 64.29 | 57.14 |
| AN | 35.71 | 42.86 | 35.71 | 42.86 | 35.71 |
| S | 35.71 | 57.14 | 35.71 | 50.00 | 35.71 |
| SL | 7.14 | 7.14 | 14.29 | 14.29 | 14.29 |

<sup>a</sup>Percent diagnostic sensitivity

SARS-CoV-2 was not detected in any complete set of paired specimens from post-symptomatic patients (**Table 3**). SARS-CoV-2 was detected in NP specimens across all platforms from 8 of 14 post-symptomatic patients. All assays and platforms detected SARS-CoV-2 in 4 AN specimens, 3 saliva specimens, and 1 SL specimen collected from post-symptomatic patients.

Considering all SARS-CoV-2-positive specimen sets regardless of symptoms, the Rheonix and EZ-SARS-CoV-2 assay performed on the ABI 7500 Fast platform detected approximately 94% of NP specimens (**Table 7**). The TaqPath COVID-19 assay detected 92% of NP specimens on the ABI 7500 Fast platform; on the QuantStudio 5 platform approximately 87 to 89% of NP specimens were detected by either real-time RT-PCR assay. Detection of AN specimens ranged from approximately 75 to nearly 81% across platforms. The EZ-SARS-CoV-2 assay was able to detect approximately 86% of saliva specimens on the ABI 7500 Fast platform and 79% on the QuantStudio 5 platform. The Rheonix platform detected approximately 78% of saliva specimens overall. The TaqPath COVID-19 assay detected 73% of saliva specimens, regardless of platform. At most, 41% of SL specimens were detected by the EZ-SARS-CoV-2 assay on the ABI 7500 Fast platform, and detection decreased to a low of 27% on the Rheonix platform (**Table 7**).

**Table 7.** Diagnostic sensitivity of Rheonix, EZ-SARS-CoV-2 (EZ), and TaqPath COVID-19 SARS-CoV-2 assays using paired specimens collected from 63 patients.

| Specimen | Rheonix | EZ<br>ABI 7500 | TaqPath<br>ABI 7500 | EZ<br>QuantStudio 5 | TaqPath<br>QuantStudio 5 |
| --- | --- | --- | --- | --- | --- |
| NP | 93.65<sup>a</sup> | 93.65 | 92.06 | 88.89 | 87.30 |
| AN | 74.60 | 80.95 | 79.37 | 79.37 | 77.78 |
| S | 77.78 | 85.71 | 73.02 | 79.37 | 73.02 |
| SL | 26.98 | 41.27 | 34.92 | 38.10 | 38.10 |

<sup>a</sup>Percent diagnostic sensitivity

The overall agreement between the 5 platforms evaluated was compiled for each specimen type, considering all 63 paired specimen sets (**Table 8**). SARS-CoV-2 detection was highest using NP specimens across platforms at 87.30%, followed by 82.54% of AN specimens, and 74.60% of saliva specimens. SARS-CoV-2 detection decreased to 61.9% in SL specimens across platforms.

**Table 8.**
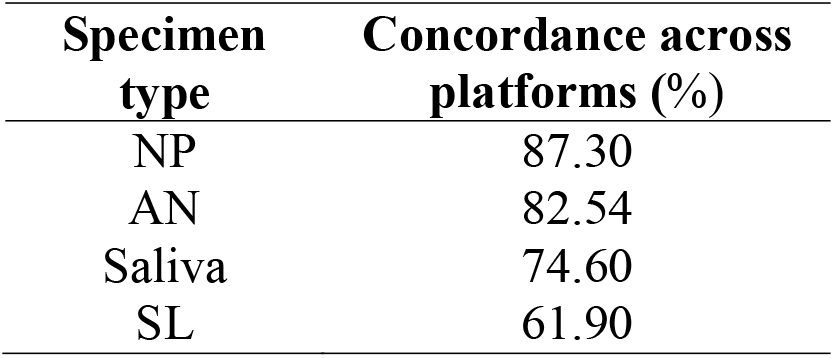
Result concordance across Rheonix, EZ-SARS-CoV-2, and TaqPath COVID-19 SARS-CoV-2 assays in paired specimen types.

| **Specimen type** | **Concordance across platforms (%)** |
| --- | --- |
| NP | 87.30 |
| AN | 82.54 |
| Saliva | 74.60 |
| SL | 61.90 |

Detection of SARS-CoV-2 RNA was most successful from NP specimens and so the percent detection relative to NP was determined for the remaining specimen types across platforms (**Table 9**). SARS-CoV-2 was detected from 79-90% of AN and saliva specimens relative to NP. However, 95% of saliva specimens were detected relative to NP when detection from all platforms were combined in contrast to 84% of AN specimens; this was largely due to positive saliva specimens collected from post-symptomatic patients (**Table 3**). Relative to NP specimens, SL specimens yielded 52% detection when results from all platforms were combined.

**Table 9.** SARS-CoV-2 detection from specimens relative to NP swabs using paired specimens collected from 63 patients.

| **Specimen** | **Rheonix** | **EZ** | **TaqPath** | **EZ** | **TaqPath** | **All platforms** |
| --- | --- | --- | --- | --- | --- | --- |
|  |  | **ABI 7500** | **ABI 7500** | **QuantStudio 5** | **QuantStudio 5** | **combined** |
| NP detected<sup>a</sup> | 59 | 59 | 58 | 56 | 55 | 62 |
| **Percent detected relative to NP** |  |  |  |  |  |  |
| AN | 79.66 | 86.44 | 84.48 | 89.29 | 89.09 | 83.87 |
| S | 81.36 | 89.83 | 79.31 | 87.50 | 83.64 | 95.16 |
| SL | 28.81 | 42.37 | 37.93 | 42.86 | 41.82 | 51.61 |

<sup>a</sup>Number of SARS-CoV-2-positive NP specimens detected

### Comparison of specimen types for detection of SARS-CoV-2

To further investigate the differences between specimen types, we compared the SARS-CoV-2 cycle threshold (Ct) value detected by the EZ-SARS-CoV-2 assay on the ABI 7500 Fast platform. Of the total 63 specimen sets, SARS-CoV-2 was detected in 59 NP specimens, 51 AN specimens, 54 saliva specimens, and 26 SL specimens (**Fig. 2**). The median Ct value in NP specimens was 26.13, the median Ct value detected in AN specimens was 26.19, and the median Ct value was 26.26 in saliva specimens. There was no difference between Ct values obtained from NP, AN, or saliva specimens (p≥0.1681). However, the median SARS-CoV-2 Ct value detected in SL specimens was zero, which indicated the lack of SARS-CoV-2 detection. SARS-CoV-2 Ct values were detected in 26 of the 63 SL specimens and in general they tend to be higher than those detected in other specimen types. SARS-CoV-2 Ct values obtained from SL specimens were different from NP, AN, and saliva specimen Ct values (p<0.0030).

**Figure 2.**
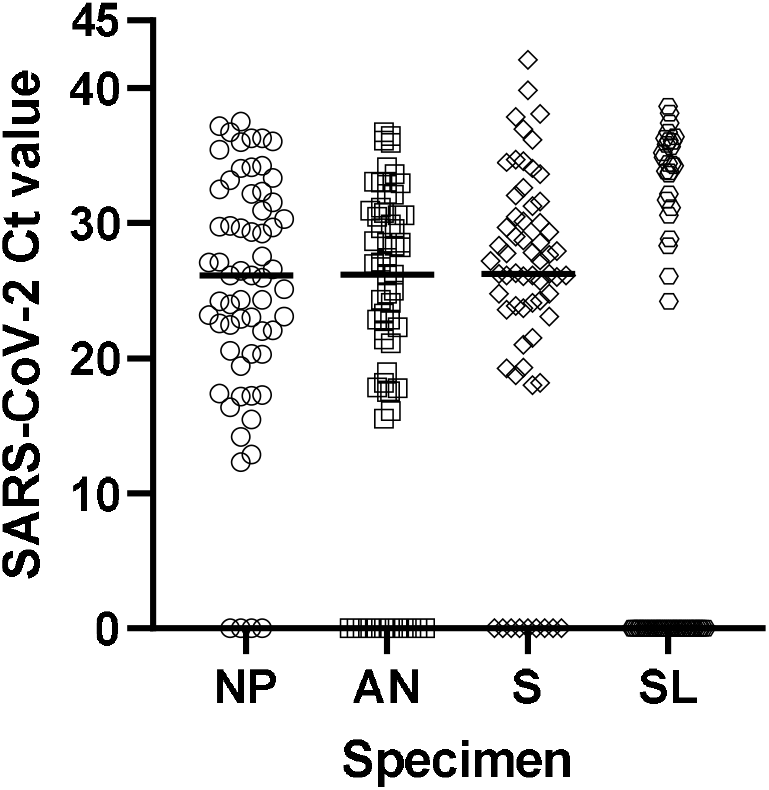
SARS-CoV-2 cycle threshold (Ct) value across paired specimen types collected from 63 positive individuals. SARS-CoV-2 cycle threshold (Ct) values obtained using the EZ-SARS-CoV-2 assay on the ABI 7500 platform are shown on the y-axis. Specimen types included nasopharyngeal swabs (NP, n=59 with positive Ct values), anterior nares swabs (AN, n=51 with positive Ct values), saliva (S, n=54 with positive Ct values), and sublingual swabs (SL, n=26 with positive Ct values), and are shown on the x-axis. The horizontal line in each specimen type indicates the median value (NP=26.13, AN=26.19, S=26.26, SL=0; 0 Ct value indicates not detected).

SARS-CoV-2 Ct values detected by the EZ-SARS-CoV-2 assay on the ABI 7500 Fast platform were also compared between symptomatic, asymptomatic, and post-symptomatic groups. No difference was found between symptomatic and asymptomatic groups for Ct values detected from NP (p=0.2307), AN (p=0.0778), saliva (p=0.2602) or SL (p=0.8490) specimens. When Ct values determined from specimens collected from post-symptomatic patients were compared to those of symptomatic or asymptomatic patients, the Ct values in the post-symptomatic group were higher for NP (p<0.0003) and saliva (p<0.0036) specimens. The difference in Ct values from post-symptomatic AN specimens neared significance when compared to symptomatic AN specimens (p=0.0581) but did not differ from asymptomatic AN specimens (p=0.7547). There were not enough Ct values from post-symptomatic SL specimens for statistical comparison.

To appreciate the dynamics of SARS-CoV-2 Ct values within a patient, only specimen sets with a Ct value for all 4 specimens from the EZ-SARS-CoV-2 assay on the ABI 7500 Fast platform were plotted (**Fig. 3**). From these 24 specimen sets, NP samples presented the lowest Ct values followed by AN (p=0.0014), S (p=0.0194), and SL (p<0.0001) samples. The latter presented the highest Ct value of all four specimen types (p<0.0001, **Fig. 3**).

**Figure 3.**
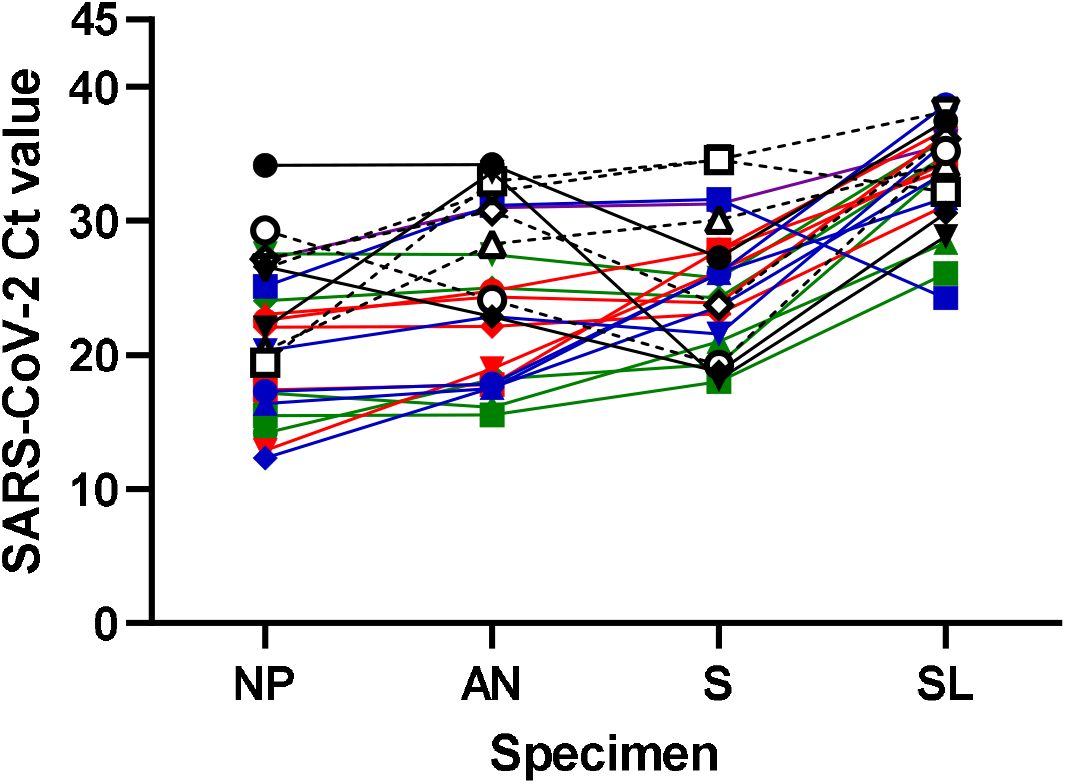
SARS-CoV-2 cycle threshold (Ct) values within sets of respiratory and oral specimens collected from 24 positive individuals. SARS-CoV-2 cycle threshold (Ct) values obtained using the EZ-SARS-CoV-2 assay on the ABI 7500 platform are shown on the y-axis. Specimen types are shown on the x-axis (NP = nasopharyngeal swab, AN = anterior nares swab, S = saliva, SL = sublingual swab). Each paired set collected from an individual patient is distinguished by a different color and symbol combination.

### SARS-CoV-2 infectivity across specimens

SARS-CoV-2 isolation was performed on 50 sets of specimens. The virus was isolated from 21 of 50 NP, 13 of 50 AN, 12 of 50 saliva, and one SL specimen (**Fig. 4**). SARS-CoV-2 isolation was most successful using specimens from symptomatic patients and isolation was not successful in specimens from post-symptomatic patients (**Fig.5**). Stratifying successful virus isolation by specimen type, NP was the specimen with the highest isolation rate, with virus isolated from 16 of 24 symptomatic and 5 of 17 asymptomatic specimens (**Fig.5A, 5B**). AN was the next most successful specimen type with virus isolated from 10 of 24 symptomatic and 3 of 17 asymptomatic specimens (**Fig.5A, 5B**). Success of virus isolation using saliva specimens was similar to AN, with 9 of 24 symptomatic and 3 of 17 asymptomatic yielding infectious virus (**Fig.5A, 5B**). Virus isolation from SL specimens resulted in only one out of 24 symptomatic and none of the 17 asymptomatic specimens yielding infectious SARS-CoV-2 when inoculated in cell culture (**Fig.5A, 5B**). SARS-CoV-2 was not isolated from post-symptomatic patient specimens (**Fig.5C**).

**Figure 4.**
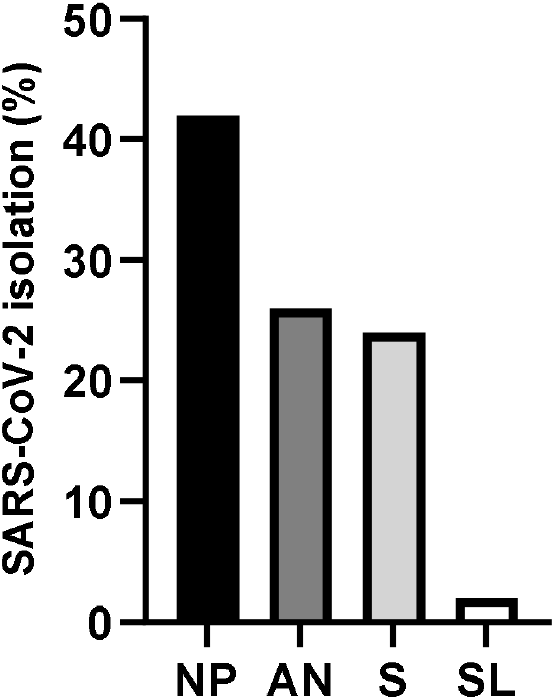
Success of SARS-CoV-2 isolation by specimen type. Specimen types are shown along the x-axis (NP = nasopharyngeal swab, AN = anterior nares swab, S = saliva, SL = sublingual swab) and the percent of specimens yielding positive SARS-CoV-2 virus isolation is shown on the y-axis.

**Figure 5.**
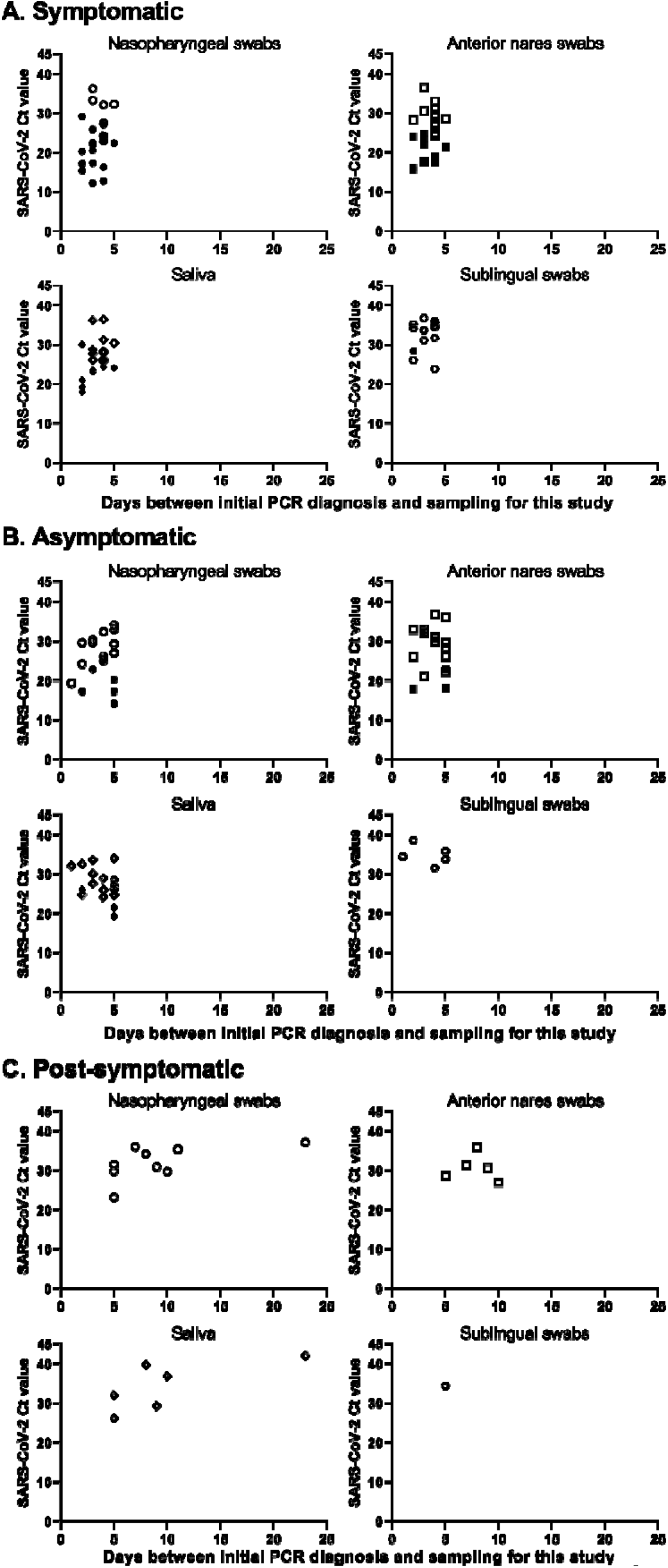
SARS-CoV-2 isolation from paired respiratory and oral specimens collected from 50 positive individuals. SARS-CoV 2 cycle threshold (Ct) values are shown on the y-axis. Days between initial diagnostic test and specimen collection for this study are on the x-axis. Successful virus isolation (filled/black symbols) or lack of virus isolation (open symbols) is

Specimens that yielded infectious SARS-CoV-2 had significantly lower Ct values for symptomatic NP (p=0.0007), AN (p<0.0001), and saliva (p=0.0009) specimens than specimens that did not yield infectious virus (**Fig.5A**). Lower SARS-CoV-2 Ct values (p≤0.0324) were also detected in asymptomatic specimens that yielded infectious virus in comparison to specimens that did not yield virus (**Fig.5B**).

The success of recovery of infectious SARS-CoV-2 decreased over time, with virus being recovered from 100% of the samples from symptomatic patients collected 2 days after initial diagnosis, which declined to 50% of the samples by 5 days (**Fig.5A**). Virus isolation was less successful from asymptomatic patient specimens, even when collected 1 day after diagnosis, although it was possible to recover SARS-CoV-2 virus from asymptomatic specimens collected up to 5 days following diagnosis (**Fig.5B**).

## Discussion

This study was undertaken to compare the performance of five molecular SARS-CoV-2 assay/platform combinations across paired respiratory (NP and AN swabs) and oral (S and SL swab) specimens collected from symptomatic, asymptomatic, and post-symptomatic patients. All five assay/platform combinations assessed in this study are based on RT-PCR. The Rheonix MDx assay incorporates cell lysis, RNA purification, amplification, and detection steps in a closed system. For the real-time RT-PCR assays, nucleic acid isolation was performed separate from amplification, although the same elution was used for all real-time assays. The limit of detection (LoD) of these assays is similar despite the differences in methodology: the Rheonix LoD is 625 genomic equivalents per mL, the EZ-SARS-CoV-2 assay LoD is 250 genome equivalents per mL and the TaqPath COVID-19 assay LoD is 10 genomic equivalents per reaction or 500 genome equivalents per mL (17, https://www.fda.gov/media/137489/download, Thermo Fisher Scientific Inc. (2020) TaqPath COVID-19 Combo Kit and TaqPath COVID-19 Combo Kit Advanced Instructions for Use, Publication Number MAN0019181, Revision H.0).

The comparative diagnostic sensitivity values determined herein revealed that the EZ-SARS-CoV-2 assay on the ABI 7500 platform was the most sensitive assay/platform combination. The Rheonix platform followed closely and demonstrated equivalent performance on NP specimens. The TaqPath COVID-19 assay on the ABI 7500 platform ranked third in performance. The QuantStudio 5 platform provided lower sensitivity, with the EZ-SARS-CoV-2 assay performing slightly better than the TaqPath COVID-19 assay.

Collection of 4 specimens from each patient allowed for direct comparisons of detection between specimens. NP specimens provided the best rate of detection among specimens collected from symptomatic and asymptomatic patients across the platforms used herein, with detection rates of 92 to 100%. NP has been described as the most sensitive specimen in other studies, even if 100% detection is not achieved (19, 20). This can likely be explained by the fact that SARS-CoV-2 replicates in nasal turbinates and the NP swab collection procedure harvests infected turbinate epithelial cells (18, 21). Detection rates of SARS-CoV-2 from AN and saliva specimens were slightly lower at 92-96% for symptomatic patients, with equivalent performance across the real-time RT-PCR platforms. However, AN and saliva specimens provided lower sensitivity when collected from asymptomatic patients (75 to 92%). A meta-analysis of published data also found a reduced detection rate from nasal swabs (AN or mid-turbinate) and saliva when compared to NP swabs collected from the same patient (20). This again, could be a result of the virus tropism and slight differences in viral loads at different replication sites (e.g. nasal turbinate epithelium vs tonsil). SARS-CoV-2 detection from sublingual swab specimens was inferior to the other specimens in this study, with detection rates of 40-60% from symptomatic patient specimens and 25 to 42% from asymptomatic patient specimens.

There was only one set of specimens collected from an asymptomatic individual (patient 33, **Table 2**) for which SARS-CoV-2 was not detected in NP on any platform; only saliva and sublingual swab specimens were positive and only when using the EZ-SARS-CoV-2 assay on the ABI 7500 platform. Detection of SARS-CoV-2 in saliva samples but not in NP has been reported for some patients across multiple studies (22–29). Senok and colleagues found that saliva specimens were especially sensitive in asymptomatic patients (30). It should be noted that the performance of saliva specimens for SARS-CoV-2 diagnosis differs according to saliva collection and processing methods (20, 23, 31). As expected and reported in the literature neither NP, nor AN or saliva specimens provide 100% sensitivity (32, 33). Some studies suggest that discordant results between paired specimens could be due to the distribution of viral replication specific to that individual (23). Additionally, the dynamics of virus infection and differences in viral load over time are also important factors that affect diagnostic sensitivity.

A strength of this study is the inclusion of symptomatic, asymptomatic, and post-symptomatic patients, allowing comparison of detection rates between these groups. The rate of detection of SARS-CoV-2 was highest using specimens from symptomatic patients (92 to 100% for NP, AN and saliva), followed by asymptomatic patient specimens (75 to 96% for NP, AN and saliva). Although detection using NP collected from post-symptomatic patients was 86% on the two most sensitive platforms, the other specimens did not robustly detect SARS-CoV-2 in post-symptomatic patients.

Equivalent SARS-CoV-2 Ct values were obtained from NP, AN, and saliva specimens when results from all 63 sets were analyzed. Comparable SARS-CoV-2 Ct values from NP and saliva specimens have also been found in other studies (27, 29, 34–37). Further, no difference was found when SARS-CoV-2 Ct values were compared between specimens collected from symptomatic and asymptomatic patients, consistent with other findings based on Ct values (38) and viral load (39). However, Ct values obtained from post-symptomatic patients NP and saliva specimens were higher than Ct values from symptomatic or asymptomatic specimens, likely reflecting a decrease in viral load after the resolution of symptoms and the infection. When the dataset was narrowed to 24 individuals for which each specimen was positive, differences in SARS-CoV-2 Ct values between specimens were observed. In this case, the SARS-CoV-2 Ct values detected from NP specimens were lower than those from AN and S, and the Ct values detected from SL specimens were higher than NP, AN, or S.

SARS-CoV-2 isolation was attempted with 50 of 63 positive sample sets and this enabled additional insights into virus infectivity among specimen types over time. Overall, SARS-CoV-2 isolation was successful in 23.5% of specimens tested. Infectious virus was isolated predominantly in specimens from symptomatic patients (16 of 25 [64%]). In contrast, infectious virus was isolated from less than one-third of specimens collected from asymptomatic patients (5 of 17 [29%]). SARS-CoV-2 viral isolation was unsuccessful when specimens from post-symptomatic patients were tested. Unfortunately, virus isolation was not possible in the original samples collected from these patients and that resulted in the first diagnosis of SARS-CoV-2 infection, as these specimens were collected in an inactivating guanidine media.

SARS-CoV-2 isolation was more successful from NP (21 of 50 [42%]) than AN (13 of 50 [26%]) or saliva (12 of 50 [24%]) specimens in this study. Specimens that yielded viral isolation were characterized by lower Ct values, corresponding to higher viral load. Lower SARS-CoV-2 Ct values and viral loads greater than 10^6^ copies per mL often contribute successful viral isolation (40–43).

Specimens collected up to 5 days after the initial SARS-CoV-2 diagnosis consistently yielded virus, although SARS-CoV-2 RNA was detected occasionally up to 23 days after diagnosis in specimens collected from post-symptomatic patients. SARS-CoV-2 shedding is known to continue up to 48 days (6, 41, 44–46). Recovery of SARS-CoV-2 by culture has been successful up to 8 days after the onset of symptoms (41, 42, 46). The continued viral RNA shedding beyond the detection of infectious virus confounds establishing guidelines for releasing individuals from isolation (47, 48). It is possible that intact viral genomes and “live” particles are present and shed, however, the viral load might be too low for successful virus isolation late in the course of infection (40, 41, 46).

Collectively, this work identified the EZ-SARS-CoV-2 assay on the ABI 7500 platform as the most sensitive SARS-CoV-2 test across patients segregated by symptomology and across specimen types. The Rheonix system also demonstrated high diagnostic sensitivity on NP, AN, and saliva specimens. Although NP specimens provided the highest sensitivity, 86-90% of paired AN and saliva specimens were detected. As AN and saliva specimens can be self-collected with minimal PPE, materials, and assistance, they offer alternatives to NP specimens. The robust detection of SARS-CoV-2 from AN and saliva specimens by the EZ-SARS-CoV-2 assay supports the use of either specimen in a surveillance program of asymptomatic individuals.

## Data Availability

All data produced in the present work are contained in the manuscript

## Acknowledgements

We thank all health care workers from Cayuga Medical Center for their help collecting NP swabs. The work was supported by the Cornell Office of the Vice President for Research.

## Notes

### Competing Interest Statement

The authors have declared no competing interest.

### Funding Statement

This study was funded by Cornell OVPR.

